# Discordance between a deep learning model and clinical-grade variant pathogenicity classification in a rare disease cohort

**DOI:** 10.1101/2024.05.22.24307756

**Authors:** Sek Won Kong, In-Hee Lee, Lauren V. Collen, Arjun K. Manrai, Scott B. Snapper, Kenneth D. Mandl

**Author notes:** Corresponding author: Sek Won Kong, MD, 401 Park Drive, LM5528.4, Boston Children’s Hospital, Boston, MA 02215.

## Abstract

Genetic testing has become an essential component in the diagnosis and management of a wide range of clinical conditions, from cancer to developmental disorders, especially in rare Mendelian diseases. Efforts to identify rare phenotype-associated variants have predominantly focused on protein-truncating variants, while the interpretation of missense variants presents a considerable challenge. Deep learning algorithms excel in various applications across biomedical tasks^1,2^, yet accurately distinguishing between pathogenic and benign genetic variants remains an elusive goal^3-5^. Specifically, even the most sophisticated models encounter difficulties in accurately assessing the pathogenicity of missense variants of uncertain significance (VUS). Our investigation of AlphaMissense (AM)^5^, the latest iteration of deep learning methods for predicting the potential functional impact of missense variants and assessing gene essentiality, reveals important limitations in its ability to identify pathogenic missense variants within a rare disease cohort. Indeed, AM struggles to accurately assess the pathogenicity of variants in intrinsically disordered regions (IDRs), leading to unreliable gene-level essentiality scores for certain genes containing IDRs. This limitation highlights the challenges in applying AM faces in the context of clinical genetics^6^.

## Introduction

Diagnostic yield for rare Mendelian diseases using whole genome and exome sequencing (WGS and WES) remains below 25%^7,8^. With 30 million genomes sequenced globally, the ever-increasing need for machine learning assistance in the interpretation of genetic variants for pathogenicity classification becomes evident. This is particularly true for the numerous missense variants that continue to be categorized as variant of uncertain significance (VUS)^9^. Despite significant advancements in deep learning methods^3-5^, current predictions often fall short of accurately predicting the pathogenicity of these mutations at scale^10^. These shortcomings are largely attributed to two factors: 1) the scarcity of gold-standard labeled datasets, and 2) the omission of genotype-phenotype associations during model training. Here, we aim to evaluate the performance and utility of the recently published deep learning model AM^5^, specifically focusing on its ability to identify pathogenic and likely pathogenic missense variants of clinical significance in well-phenotyped individuals within a rare disease cohort^11,12^.

## Results

First, we examine agreement between missense variants identified as likely pathogenic by AM (AM_LP) and those expertly curated as pathogenic and likely pathogenic in ClinVar (ClinVar_P and ClinVar_LP, respectively)^9^, within a cohort of 7,454 individuals with rare diseases and their family members. This cohort comprises 3,383 patients with rare diseases presumed to be of genetic origin, along with their family members. By comparing AM’s classification of missense variants with expert-curated data from ClinVar^9^, we estimate the rates of false positives and false negatives. This analysis addresses the disparity between the balanced distribution of pathogenic and benign variants in gold-standard datasets, which are used for algorithm fine-tuning, and the rarer occurrence of pathogenic variants in real-world datasets.

Subsequently, we conducted a comparative analysis of AM’s performance against other deep-learning approaches, such as ESM1b^4^ and EVE^3^, as well as against an established non-deep learning method, the rare exome variant ensemble learner (REVEL)^13^. ESM1b and EVE were selected for comparison with AM because they utilize deep learning algorithms and demonstrated similar top performances in Cheung *et al*.’s study^5^. EVE employs an unsupervised generative autoencoder, showcasing superior performance for a limited set of well-aligned proteins and residues. In contrast, ESM1b covers entire protein-coding genes by pre-training protein language models with samples across all organisms, thus providing scores for regions not covered by multiple sequencing alignments. REVEL was selected for comparison with AM because it has been integrated into gene-specific American College of Medical Genetics and Genomics (ACMG) guidelines developed by various Variant Curation Expert Panels within ClinGen^14-16^. Coverage across human protein-coding genes varies among methods as per dbNFSP v4.6: EVE assesses 2,895 genes, ESM1b evaluates 18,884 genes, REVEL covers 18,289 genes, and AM includes 18,982 genes (**Fig 1a**). Excluding EVE, which has smaller coverage, ESM1b, REVEL, and AM covers 16,708 genes with variants reported in ClinVar in common (**Supplementary Fig. 1**). Our analysis, focused on evaluating AM’s precision, is constrained by the intersection of AM’s gene coverage and expertly curated variants in ClinVar, opting not to undertake a comprehensive benchmark across all deep learning methods.

**Fig. 1.**
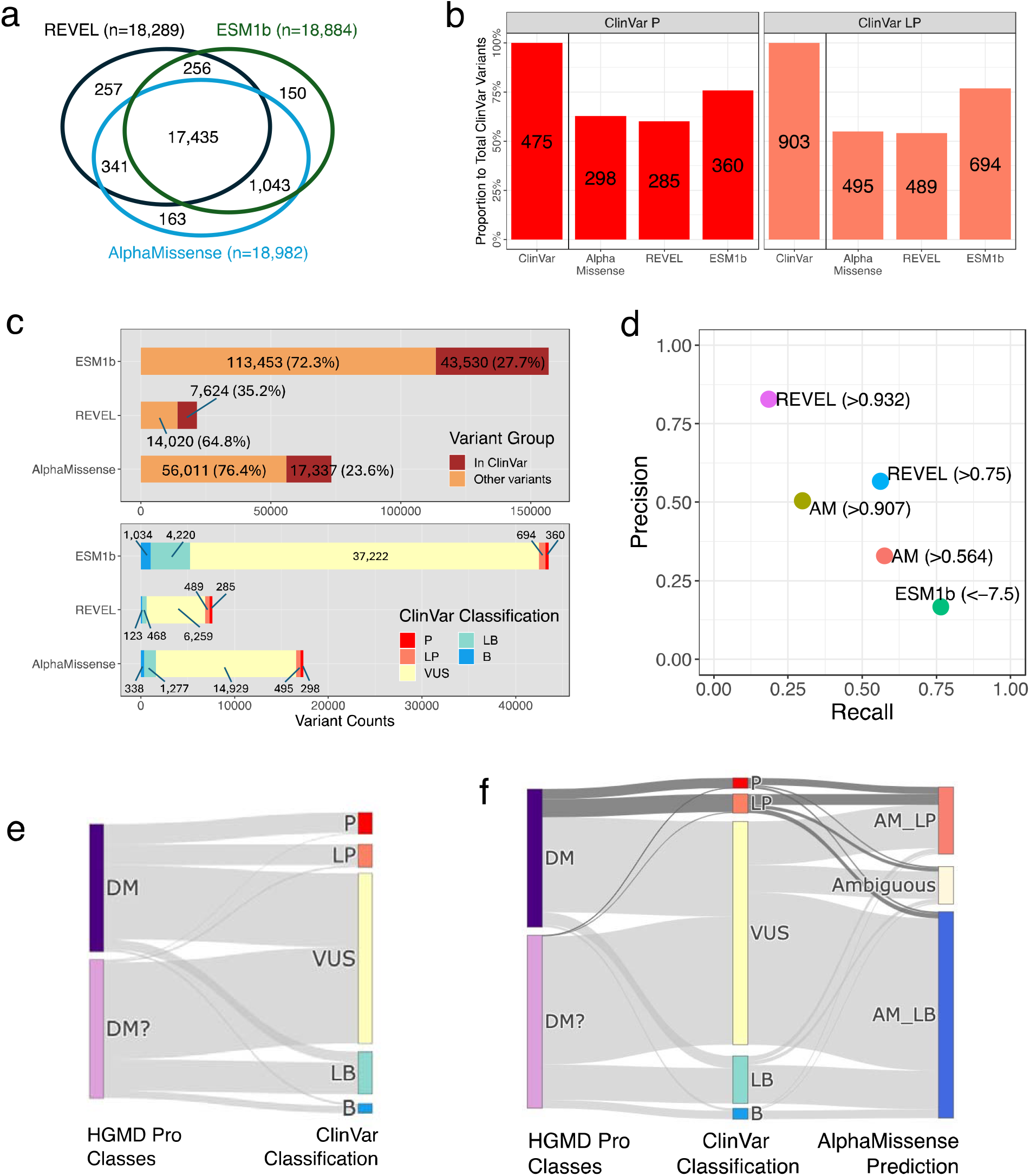
Evaluation of AlphaMissense compared to expert-curated genotype-phenotype association databases within a rare disease cohort. **a**. Gene coverage comparison across missense variant effect prediction algorithms. **b**. Counts of likely pathogenic variants predicted by AlphaMissense, REVEL, and ESM1b among ClinVar pathogenic (ClinVar_P) and likely pathogenic (ClinVar_LP) variants in a rare disease cohort. The y-axis represents the proportion of the total ClinVar_P or ClinVar_LP varaints found in the cohort. The numbers show the variant counts in each category and algorithm. **c**. The counts and proportions of variants reported in ClinVar among total variants classified as likely pathogenic in each algorithm (top panel). The stratification of variants that were both classified as likely pathogenic in each algorithm and reported in ClinVar, according to the classification by ClinVar (bottom panel). **d**. Precision and recall for each algorithm. AM: AlphaMissense. **e**. Concordance between ClinVar’s pathogenic or likely pathogenic classifications and HGMD’s DM and DM? classes. **f**. Discrepancies between HGMD Pro (DM and DM? classes) and ClinVar classification/AlphaMissense prediction in our cohort.

Identifying phenotype-associated variants in individuals with rare diseases is a two-step process: first, by predicting and classifying variant effects, and subsequently, by prioritizing these variants within genes linked to presenting phenotypes^17^(**Supplementary Fig.2**). The lack of phenotype information in the development (or fine-tuning) of AM and other prediction algorithms required a comparison of AM’s pathogenicity scores with ClinVar’s pathogenicity classifications within our well-phenotyped patient cohort. AM identified 298 of 475 ClinVar_P (62.7%) and 495 of 903 ClinVar_LP (54.8%) variants were classified as AM_LP **(Fig. 1b)**. Conversely, 17,337 (23.6% of total rare variants classified as AM_LP, n=73,348, gnomAD minor allele frequency < 1%) variants were also reported in ClinVar, regardless of reported pathogenicity (**Fig. 1c top panel**). Of these, only 298 (1.7% of 17,337) and 495 (2.9% of 17,337) were annotated as ClinVar_P and ClinVar_LP, respectively (**Fig. 1c bottom panel**). In contrast, REVEL, a non-deep learning but formerly cutting-edge method, identified 285 (3.7% of 7,624) of ClinVar_P and 489 (6.4% of 7,624) of ClinVar_LP variants with a threshold above 0.75. Precision and recall for rare missense variants discovered in our cohort and also curated in ClinVar for AM, REVEL and ESM1b are detailed in **Fig. 1d** and **Supplementary Tab. 1**. AM’s performance on reported ClinVar_P and _LP variants demonstrated a preference for recall over precision.

After assessing AM’s classifications against the current gold standard, ClinVar, and other algorithms, we expanded our analysis to include discrepancies among expert-curated databases as well as their alignment with AM within our cohort. The comparison of the disease-causing mutation (DM) and likely disease-causing mutation (DM?) categories from the Human Gene Mutation Database (HGMD) Professional^18^ (release Q1-2023) with ClinVar’s annotations showed limited agreement^19^. Specifically, the DM and DM? categories from HGMD aligned with ClinVar’s P or LP classification at rates of 27.0% and 1.5%, respectively (**Fig. 1e**). In contrast, AM classified 55.8% of DM and 74.4% of DM? classifications from HGMD as likely benign (AM_LB), most of which were classified as VUS, LB, or B in ClinVar. Despite these differences, the majority of variants classified as ClinVar_P and _LP were closely aligned with HGMD’s DM and DM? categories and classified as likely pathogenic (AM_LP) by AM (**Fig. 1f**). These observations highlight the critical importance of expert consensus in accurately classifying variant pathogenicity and shed light on the complexities involved in interpreting missense variants for rare diseases. Moreover, our findings indicate that AM, while advanced, needs further development to effectively tackle these challenges.

Next, we concentrated on rare diseases with consensus candidate genes to demonstrate the practical utility of AM in prioritizing clinically meaningful genetic variants. We assessed AM’s performance within a cohort diagnosed with inflammatory bowel disease (IBD), specifically those with very early onset (less than 6 years of age at IBD onset) and early onset (less than 10 years of age at IBD onset). This population was selected based on the increased likelihood of monogenic disease (i.e., monogenic IBD (mIBD)) and the availability of expert-curated candidate genes for mIBD^20^. Subjects are a prospectively enrolled, broadly consented, WES sequenced cohort of 750 patients with IBD at a large pediatric teaching hospital (**Online Methods**)^11^. All genomic variants with allele frequency below 1% in gnomAD were selected for further evaluation, as mIBD is considered as a rare condition. Also, considering AM’s preference for recall over precision, we calibrated the threshold to categorizing variants as likely pathogenic to have high estimated positive predictive value using methods suggested by ClinGen Sequence Variant Interpretation Working Group^6^. For AM_LP, a threshold of 0.907 was used to achieve 0.98 of posterior probability of pathogenicity (described in the **online Methods** section). For each individual diagnosed with IBD, we prioritized potentially disease-associated genetic variants within a consensus list of 102 mIBD genes (**Supplementary Tab. 2**), in the order of loss-of-function (LoF), ClinVar_P, ClinVar_LP, and AM_LP (**Fig. 2a left panel, Supplementary Tab. 3**)^21^. We identified LoF variants in 77 (10.3% of 750) patients and ClinVar_P or _LP variants in 34 (4.5% of 750) patients, including 16 with LoF variants as well. In total, 95 patients (12.7% of 750) possessed genetic variants that require further validation and interpretation, before considering AM_LP variants.

**Fig. 2.**
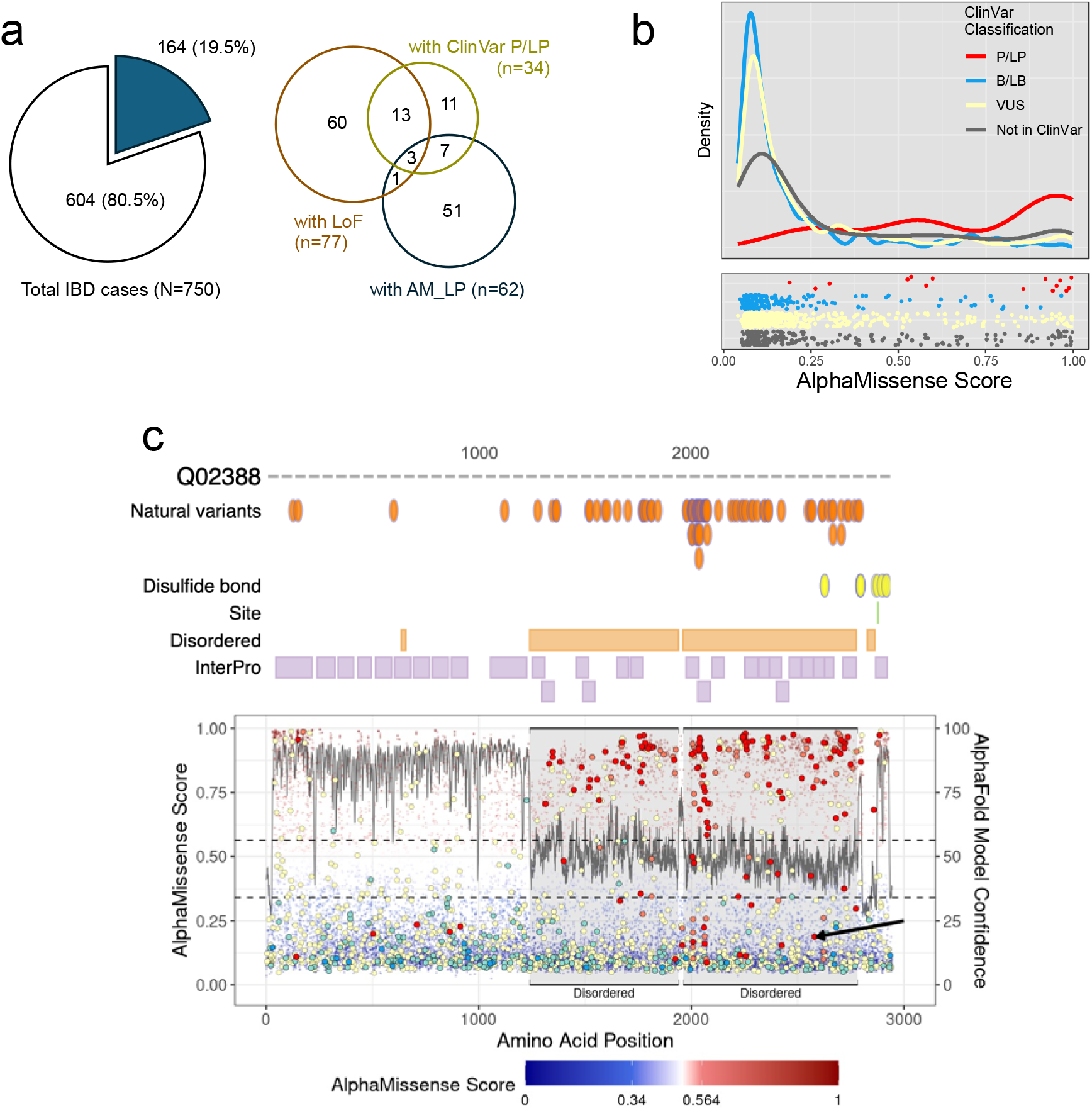
ClinVar classification and AlphaMissense pathogenicity scores for the variants in individuals with inflammatory bowel diseases in our rare disease cohort. **a**. The number and proportion of patients, out of the total cohort with inflammatory bowel diseases (IBD), with loss-of-function (LoF), ClinVar pathogenic (ClinVar_P) or likely pathogenic (ClinVar_LP), or likely pathogenic by AlphaMissense (left). The composition of patients according to the combination of variants found in each patient (right). **b**. The distribution of pathogenicity scores by AlphaMissense for ClinVar variants and other missense variants in candidate genes. The vertical line represents the thresholds for likely benign (0.34) and likely pathogenic (0.564) for AlphaMissense. **c**. Experimentally validated protein domain structure of COL7A1 (UniProt accession number: Q02388) (top panel). The known disordered region in the COL7A1 protein is marked by two orange boxes. Distribution of AlphaMissense pathogenicity scores along amino acid positions with reported ClinVar pathogenic (red circles) variants (bottom panel). The reported pathogenic variants are frequently misclassified as likely benign by AlphaMissense, especially in the known disordered region (grey highlight, marked by AlphaFold2 model confidence < 50). The variant highlighted with an arrow shows the misclassified pathogenic variant found in our cohort.

A total of 62 patients (8.3% of 750) had a median of one AM_LP variants (range 1-2) within the 102 candidate genes, with various combination of LoF or ClinVar variants (**Fig. 2a right panel, Supplementary Tab. 4**). Of the 48 variants found in these 62 patients, five (10.4%) had findings also supported by ClinVar as P or LP. Except for the 20 variants that were not reported in ClinVar, the rest of AM_LP variants were either reported as VUS (n=20), ClinVar_B (n=1), or reported as pathogenic but without criteria specified (n=2). Out of the 62 patients, 4 had both LoF and AM_LP variants within the candidate genes. An interesting case to illustrate involved a patient with congenital enteropathy, ocular and gonadal abnormalities, who exhibited compound heterozygous variant in the *WNT2B* gene. The maternally inherited frameshift variant was reported as VUS in ClinVar. The presumably paternally inherited missense variant, NM_024494.3(WNT2B):c.722G>A (p.Gly241Asp) was classified as AM_LP (**Supplementary Fig. 3**). Indeed, all computational algorithms–AM, ESM1b, and REVEL–predicted this missense variant as likely pathogenic. This case, along with two others, has been reported as part of an oculo-intestinal syndrome attributed to genetic variants in the *WNT2B* gene^22^. In clinical genetics practice, LoF variants are generally prioritized over missense variants, except when the same amino acid change has already been established as pathogenic mechanism^23^. Excluding the 4 cases with LoF variants and the 7 cases with ClinVar_P/LP variants, there were 51 cases (6.8% of 750, **Fig. 2a right panel**) presented AM_LP variants, averaging one variant per case within the same set of 102 candidate genes, which could be considered for further evaluation depending on their zygosity. This suggests the potential utility of AM’s pathogenicity scores in elucidating the putative genetic basis for additional cases. Nonetheless, functional studies and additional evaluations are crucial to confirm the pathogenicity of these variants for mIBD. Upon closer examination of the ClinVar_P/LP variants identified in IBD cases, some failed to meet the threshold for classification as AM_LP within the 102 candidate genes, indicating false negatives (**Fig.2b, Supplementary Tab. 4**). There were 13 missense variants classified as P or LP in ClinVar, eight of which were scored below the threshold of 0.907, including five with scores less than 0.564, the threshold used in original release of prediction results by AM^5^.

**Fig. 3.**
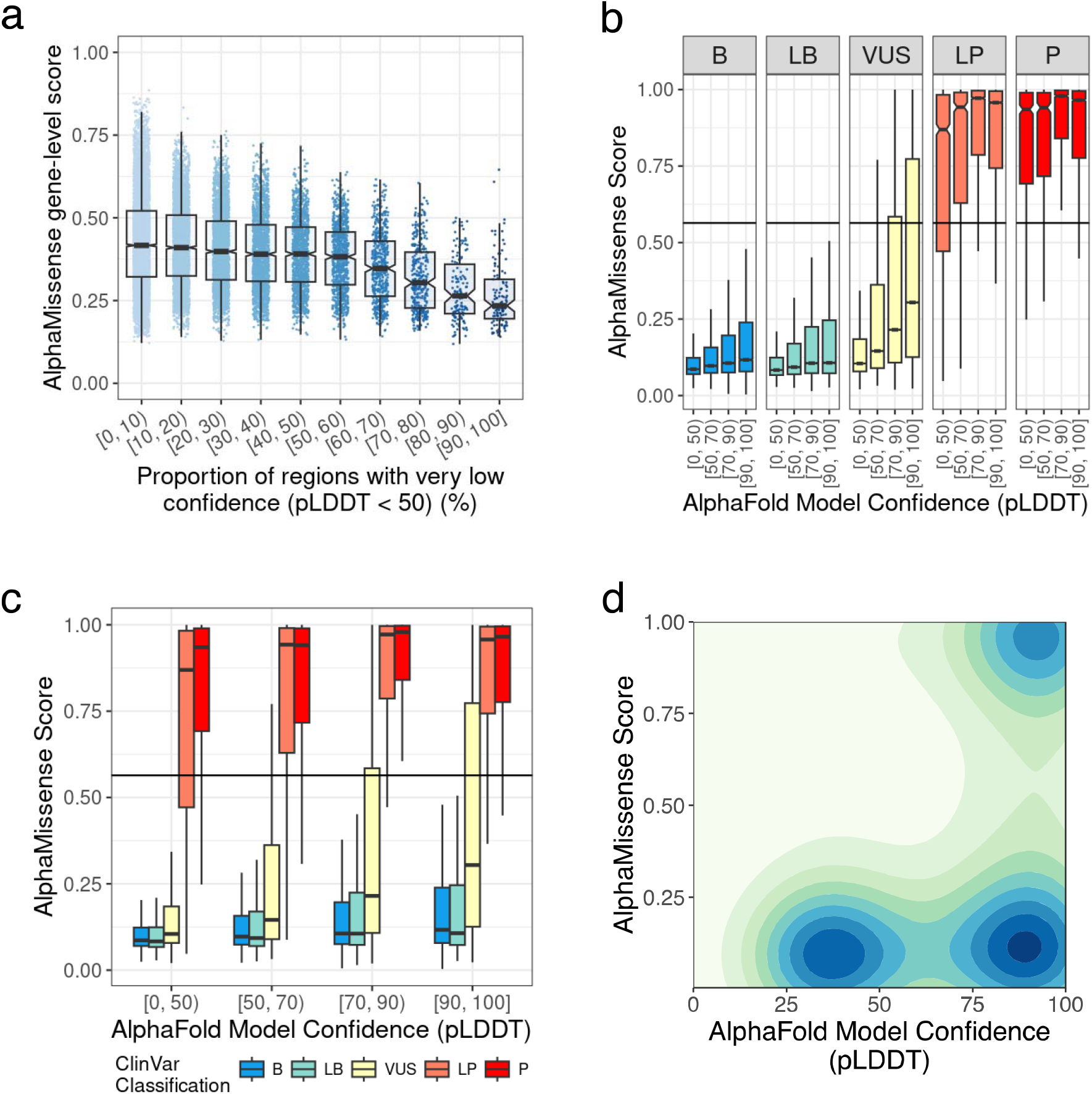
AlphaMissense pathogenicity scores and per-residue model confidence scores of AlphaFold2. **a**. AlphaMissense (AM) gene-level scores, derived from averaging all AM pathogenicity scores per gene, show a negative correlation with regions of very low model confidence in AlphaFold2 (per-residue model confidence scores (pLDDT) < 50). **b**. A negative correlation between AlphaFold2 model confidence and AM pathogenicity scores. The horizontal line represents the threshold for likely pathogenic (0.564) for AlphaMissense. **c**. Discrepancy between ClinVar P/LP classifications and AM predictions in regions with very low AlphaFold2 model confidence (pLDDT < 50). The horizontal line represents the threshold for likely pathogenic (0.564) for AlphaMissense. **d**. 2D density plot of AM scores for all ClinVar variants.

Intrinsically disordered regions (IDRs) are protein segments lacking an ordered three-dimensional structure, facilitating versatile molecular interactions^24^. Interestingly, some of the discordant variants between ClinVar and AM were found to be located within IDRs^25^. For instance, the *COL7A1* gene, encoding type VII collagen (UniProt accession number: Q02388), comprises of 2,944 amino acids (aa), where two non-collagenous domains (1-1,240 aa and 2,880-2,930 aa) flank a collagenous domain (1,240-2,880 aa) (**Fig.2c top panel**). Notably, the collagenous domain includes an intrinsically disordered hinge region (indicated as disordered in **Fig. 2c**). IDRs typically show the per-residue model confidence score (pLDDT), a measure of local accuracy according to AlphaFold2, below 50, as shown with the grey line in **Fig. 2c bottom panel**^26^. Indeed, the same region showed increased number of variants where the ClinVar_P/LP variants were not classified as AM_LP (**Fig 2c bottom panel**).

Considering that AlphaFold2’s predicted three-dimensional structures were used to pre-train AM’s deep learning models, we suspected that regions predicted with low confidence by AlphaFold2 would correspond to areas with generally lower AM pathogenicity scores, leading to discordance with ClinVar’s P/LP classifications. For this analysis, we examined all protein coding genes with AM pathogenicity scores (n=19,084). AM’s gene-level scores are calculated by averaging all possible AM pathogenicity scores for each gene. Indeed, AM gene-level scores were negatively correlated with the proportion of very low confidence (pLDDT < 50) (**Fig.3a**). At the variant level, AM pathogenicity scores exhibited a negative correlation with pLDDT scores (**Fig. 3b**), and discrepancy between ClinVar_P/LP and AM_LP classifications was significantly greater for variants found in regions with pLDDT < 50 (**Fig. 3c**). Furthermore, the variants in regions with pLDDT scores below 50 rarely scored highly by AM regardless of their classifications in ClinVar (**Fig. 3d** and **Supplementary Fig. 4**), leading to increased discrepancies with ClinVar classifications and inaccurate gene-level essentiality scores. These findings underscore the challenges AM faces in accurately predicting the impact of missense variants in certain regions and genes.

## Discussion

Deep learning models demonstrate state-of-the-art performance across a wide range of biomedical fields^1,2^. However, hasty adoption of such models could pose risks, as exemplified by the failure of a sepsis prediction algorithm, which produced a substantial number of false positives and false negatives within an electronic health record system^27^. This underscores the critical need for thorough evaluation before integrating deep learning-based algorithms into clinical and research workflows^28^. In summary, we found that there was significant discordance between pathogenicity predictions derived from a novel deep learning model and those provided by ClinVar, particularly for variants located in IDRs and for intrinsically disordered proteins (IDPs). Furthermore, clinical genetics prioritizes precision over recall due to the challenge of assessing numerous genomic variants per individual^29^. In contrast, computational algorithms tend to prioritize recall over precision, resulting a larger number of false positives when identifying genetic variants in individuals with rare diseases without knowledge of candidate genes. This study is limited by its partial focus on a disease cohort from a single center and relying solely on ClinVar for assessing the clinical significance of variants. The integration of deep learning algorithms into clinical genetics workflows requires careful evaluation of potentials for false positive and false negative findings. Comprehensive studies that integrate detailed phenotype information with genotype data are crucial for advancing prediction algorithms beyond the current capabilities of ClinVar, thereby enhancing the application of these algorithms in clinical genetics.

## Methods

The present study was approved by the Boston Children’s Hospital Institutional Review Board under protocol number P00000159.

### A cohort of individuals diagnosed with rare diseases

The analysis focuses on a cohort of 7,454 individuals, comprising 3,383 individuals diagnosed with a spectrum of rare diseases, along with their family members. This group was enrolled through the efforts of 51 clinicians at Boston Children’s Hospital, motivated by clinical presentations that hinted at genetic underpinnings. For comprehensive details on the project investigators, the main reasons for enrollment, and the genetic testing methods employed, refer to the Supplementary Table. Clinicians recorded phenotypes in RedCap, and the human phenotype ontology (HPO) terms were aligned with electronic health record extracts by CliniThink. The analysis utilized HPO terms and the primary diagnoses made by clinicians. The three most common diagnoses within the cohort were epilepsy (n=1,723), inflammatory bowel disease (IBD) (n=1,430), and congenital sensory neural hearing loss (n=842).

### WES dataset and annotation of genomic variants

Whole-exome sequencing (WES) data were uniformly generated by a single vendor, utilizing the same exome capture kit across all samples. The processing of the WES data was carried out on the DRAGEN Bio-IT Platform germline workflow (v3.9). A merged variant call file (VCF) served as the basis for the current study. These variants were then annotated using the Ensembl Variant Effect Predictor (VEP) release 110 to calculate functional consequences on for 61,552 Ensembl genes, along with variant allele frequencies from gnomAD database (release 3.0, gnomAD genomes). In addition to functional consequences, we extracted clinical significances, review statuses, and the disease name for each variant in ClinVar (ClinVar 20231209) and mutation category for the variants in Human Gene Mutation Database (HGMD Professional, Q1-2023 release) to annotate variants in WES data. Finally, for each missense variant found in WES data, we extracted the pathogenicity scores for the matching variant and transcript from AlphaMissense (available at https://zenodo.org/records/8208688, scores of the variants in hg38 coordinates for 19,233 canonical transcripts only) and ESM1b (available at https://huggingface.co/spaces/ntranoslab/esm_variants, scores of all possible single amino acid change for 42,286 human isoforms). For pathogenicity scores from REVEL, we extracted scores compiled in dbNSFP v4.6.

### Prioritizing variants for further clinical evaluation in inflammatory bowel disease cohort

For analysis of inflammatory bowel disease cases, candidate variants were selected by the following two steps. First, as monogenic inflammatory bowel disease is a rare condition, we only considered variants with a variant allele frequency of 1% or less in gnomAD genomes (version 3) in any of five population group as identified among gnomAD genomes: nfe (non-Finnish European), amr (Admixed American), eas (East Asian), sas (South Asian), and afr (African/African American). Then, we selected (1) variants whose functional consequences can be considered as loss of protein function (LoF), (2) variants which were reported as pathogenic or likely pathogenic in ClinVar (ClinVar 20231209), or (3) missense variants which were classified as ‘likely pathogenic’ by AlphaMissense (AM_LP).

Specifically, variants were considered as LoF if their functional consequences were one of the following terms in Sequence Ontology: frameshift_variant, splice_donor_variant, splice_acceptor_variant, stop_gained, stop_lost, and start_lost. The variants reported in ClinVar were further filtered by the quality of supporting evidence in their review status (https://www.ncbi.nlm.nih.gov/clinvar/docs/review_status/). In this study, only variants with at least one gold star or more – variants submitted with assertion criteria and evidence – were considered. Finally, for pathogenicity prediction by AlphaMissense, the threshold to classify variants as likely pathogenic was adjusted using the computational framework for missense variant pathogenicity classification proposed by ClinGen Working Group^6^. Here, the thresholds for pathogenicity scores were iteratively searched to achieve posterior probability for pathogenicity (or benignity) according to the desired strength of evidence (supporting, moderate, strong, or very strong), using a subset of ClinVar variants. The threshold value of 0.907 for AM_LP variants was selected to achieve very strong evidence for pathogenicity (corresponding to the posterior probability of pathogenicity of 0.98).

## Data Availability

The individual-level phenotype and genotype data used in this study is available to approved researchers via Genomic Information Commons portal at: https://pl-gic.childrens.harvard.edu/.

## Code availability

Variant annotation was performed using VEP release 110. Compilation of pathogenicity scores by AlphaMissense, ESM1b, and REVEL was performed using R programming language (v4.1.3). The code is available from the author (I.-H. L.) on request.

## Author Contributions

Dr. Kong had full access to all of the data in the study and take responsibility for the integrity of the data and the accuracy of the data analysis.

Concept and design: Kong, Manrai, Mandl.

Acquisition, analysis, or interpretation of data: Kong, Lee, Collen, Snapper.

Drafting of the manuscript: Kong, Lee, Collen, Mandl.

Critical review of the manuscript for important intellectual content: All authors.

Obtained funding: Kong, Snapper, Mandl

Administrative, technical, or material support: Kong, Lee, Collen, Snapper, Mandl

Supervision: Kong

