## Supplementary Tables for "Discordance between a deep learning model and clinical-grade variant pathogenicity classification in a rare disease cohort"

Supplementary Table 1. Precision and recall for AlphaMissense, ESM1b, and REVEL using rare missense variants discovered in our rare disease cohort and also curated in ClinVar.
Supplementary Table 2. List of 102 mIBD candidate genes used in the analysis.
Supplmenetary Table 3. List of rare (gnomAD AF < 1%) loss-of-function, ClinVar P/LP variants in 102 mIBD candidate genes, observed in 750 cases (restricted).
Supplementary Table 4. List of rare (gnomAD AF < 1%) AM-LP variants in 102 mIBD candidate genes, observed in 750 cases (restricted).

Supplementary Table 1. Precision and recall for AlphaMissense, ESM1b, and REVEL using rare missense variants discovered in our rare disease cohort and also curated in ClinVar.

| **Algorithm Name** | **Precision** | **Recall** |
|---|---|---|
| AlphaMissense | | |
| - Likely pathogenic if score > 0.564 | 0.3293 | 0.5755 |
| - Likely pathogenic if score > 0.907 | 0.5043 | 0.2983 |
| ESM1b | | |
| - Likely pathogenic if score < -7.5 | 0.1671 | 0.7649 |
| REVEL | | |
| - Likely pathogenic if score > 0.75 | 0.567 | 0.5617 |
| - Likely pathogenic if score > 0.932 | 0.8285 | 0.1858 |

### Supplementary Table 2. List of 102 mIBD candidate genes used in the analysis.

Reference: An Integrated Taxonomy for Monogenic Inflammatory Bowel Disease, Bolton *et al.* , *Gastroenterology* **162**:859-876, 2022

| Gene Name |
|---|
| ADA |
| ADA2 |
| AGR2 |
| ALPI |
| ANKZF1 |
| ARPC1B |
| BACH2 |
| BCL10 |
| BTK |
| CARD11 |
| CARD8 |
| CARMIL2 |
| CASP8 |
| CD3G |
| CD40LG |
| CD55 |
| CDC42 |
| COL7A1 |
| CTLA4 |
| CYBA |
| CYBB |
| CYBC1 |
| DCLRE1C |
| DKC1 |
| DOCK2 |
| ELF4 |
| FERMT1 |
| FMNL2 |
| FOXP3 |
| G6PC3 |
| GUCY2C |
| HPS1 |
| HPS3 |
| HPS4 |
| HSPA1L |
| ICOS |
| IKBKG |
| IKZF1 |
| IL10 |
| IL10RA |
| IL10RB |
| IL21 |
| IL2RA |
| IL2RB |
| IL2RG |
| IL37 |
| IRF2BP2 |
| ITCH |
| ITGB2 |
| LACC1 |
| LIG4 |
| LRBA |
| MALT1 |
| MASP2 |
| MVK |
| NCF1 |
| NCF2 |
| NCF4 |
| NFAT5 |
| NFKBA |
| NLRC4 |
| NOP10 |
| NPC1 |
| PI4KA |
| PIK3CD |
| PIK3R1 |
| POLA1 |
| PRKCD |
| PTPN2 |
| RAG1 |
| RAG2 |
| RBCK1 |
| RELA |
| RIPK1 |
| RTEL1 |
| SAMD9 |
| SCGN |
| SIRT1 |
| SKIC2 |
| SKIC3 |
| SLC26A3 |
| SLC37A4 |
| SLC9A3 |
| SLCO2A1 |
| STAT1 |
| STAT3 |
| STIM1 |
| STXBP3 |
| SYK |
| TGFB1 |
| TGFBR1 |
| TGFBR2 |
| TNFAIP3 |
| TRIM22 |
| TRNT1 |
| TTC7A |
| TYMP |
| WAS |
| WIPF1 |
| WNT2B |
| XIAP |
| ZAP70 |
