## Supplementary Figures for "Discordance between a deep learning model and clinical-grade variant pathogenicity classification in a rare disease cohort"

### Slide 1
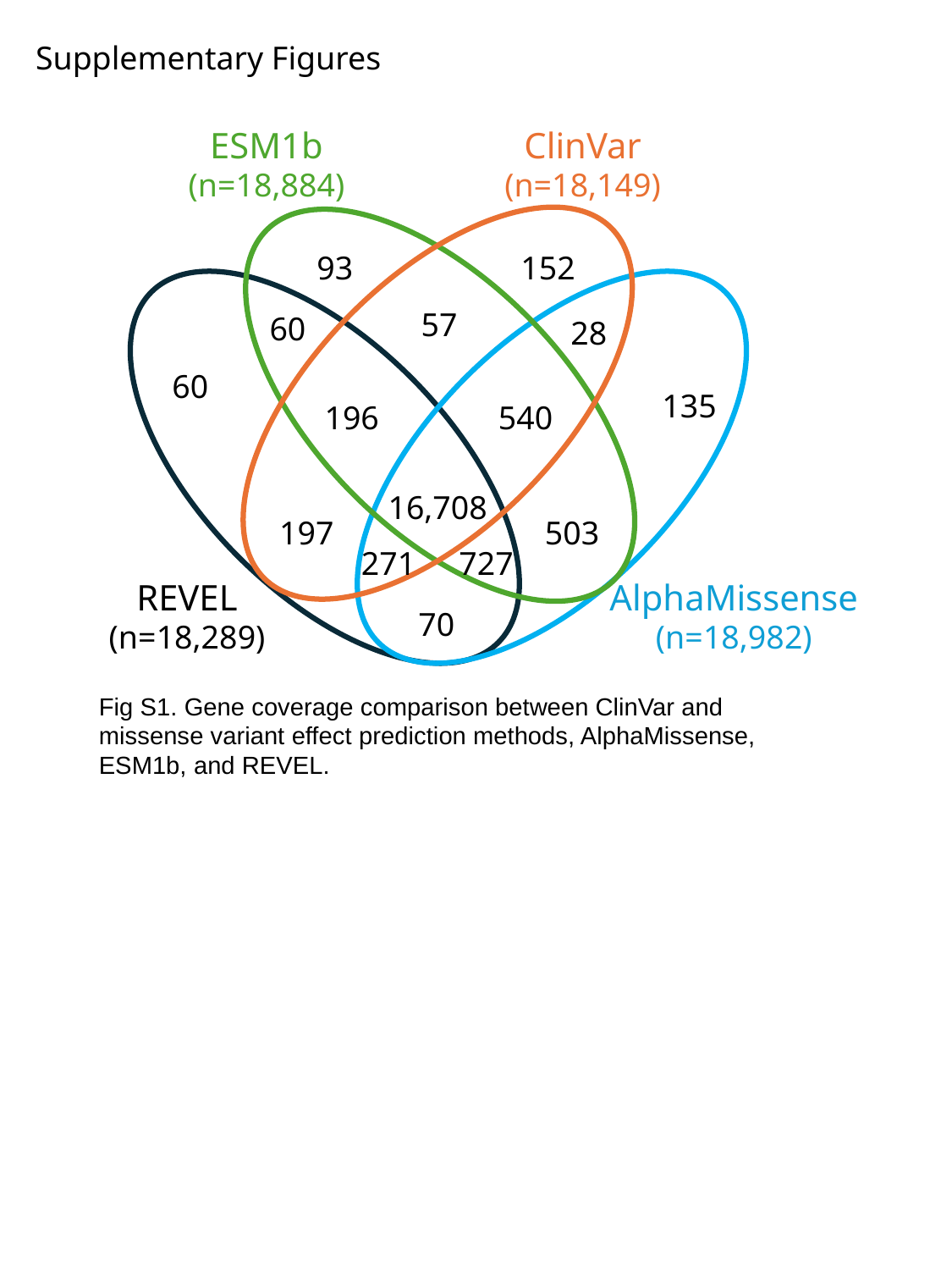

Supplementary Figures
ESM1b
(n=18,884)
ClinVar
(n=18,149)
93
152
57
60
28
60
135
196
540
16,708
197
503
271
727
REVEL
(n=18,289)
AlphaMissense
(n=18,982)
70
Fig S1. Gene coverage comparison between ClinVar and missense variant effect prediction methods, AlphaMissense, ESM1b, and REVEL.

### Slide 2
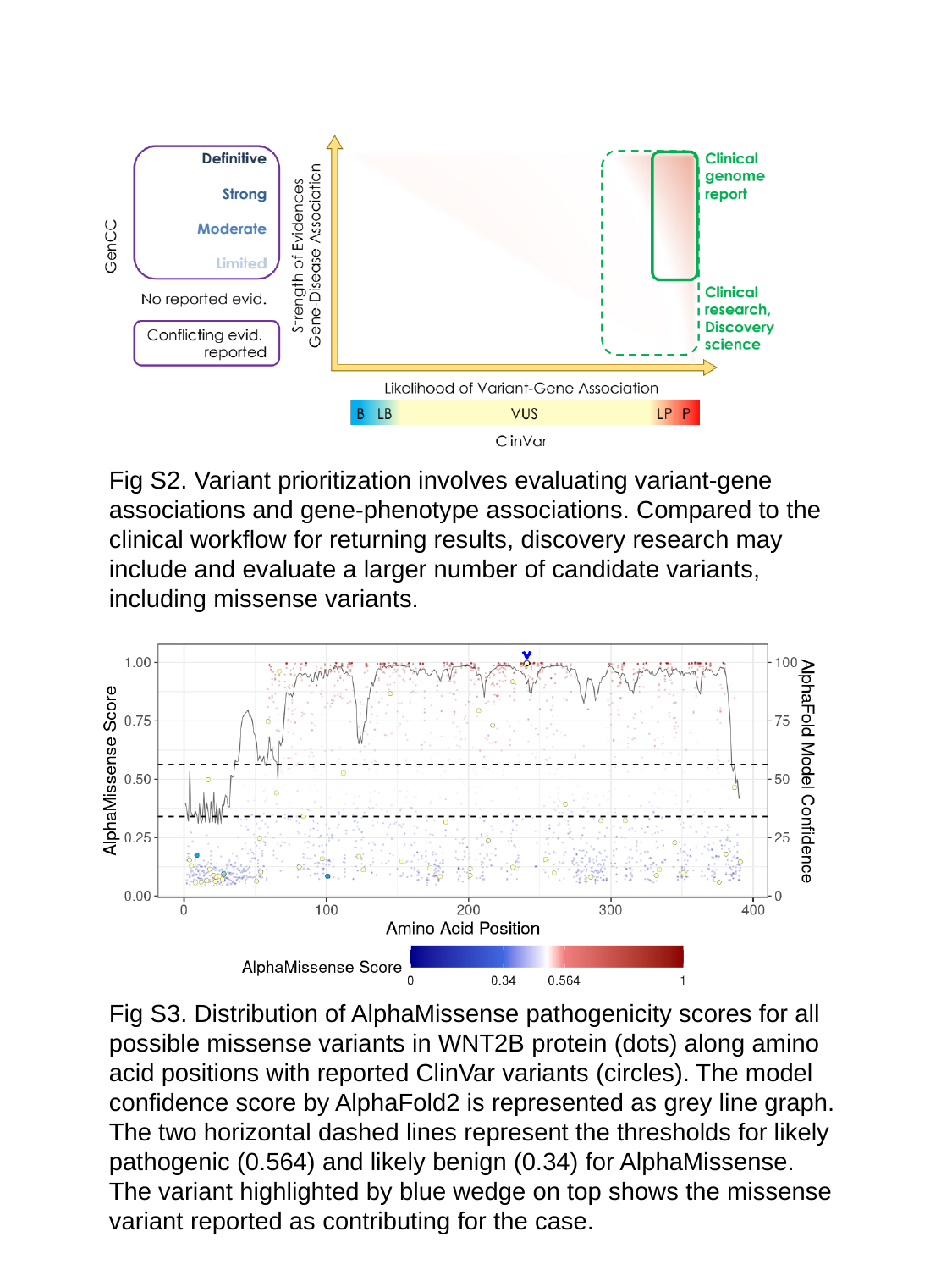

Fig S2. Variant prioritization involves evaluating variant-gene associations and gene-phenotype associations. Compared to the clinical workflow for returning results, discovery research may include and evaluate a larger number of candidate variants, including missense variants.
Fig S3. Distribution of AlphaMissense pathogenicity scores for all possible missense variants in WNT2B protein (dots) along amino acid positions with reported ClinVar variants (circles). The model confidence score by AlphaFold2 is represented as grey line graph. The two horizontal dashed lines represent the thresholds for likely pathogenic (0.564) and likely benign (0.34) for AlphaMissense. The variant highlighted by blue wedge on top shows the missense variant reported as contributing for the case.

### Slide 3
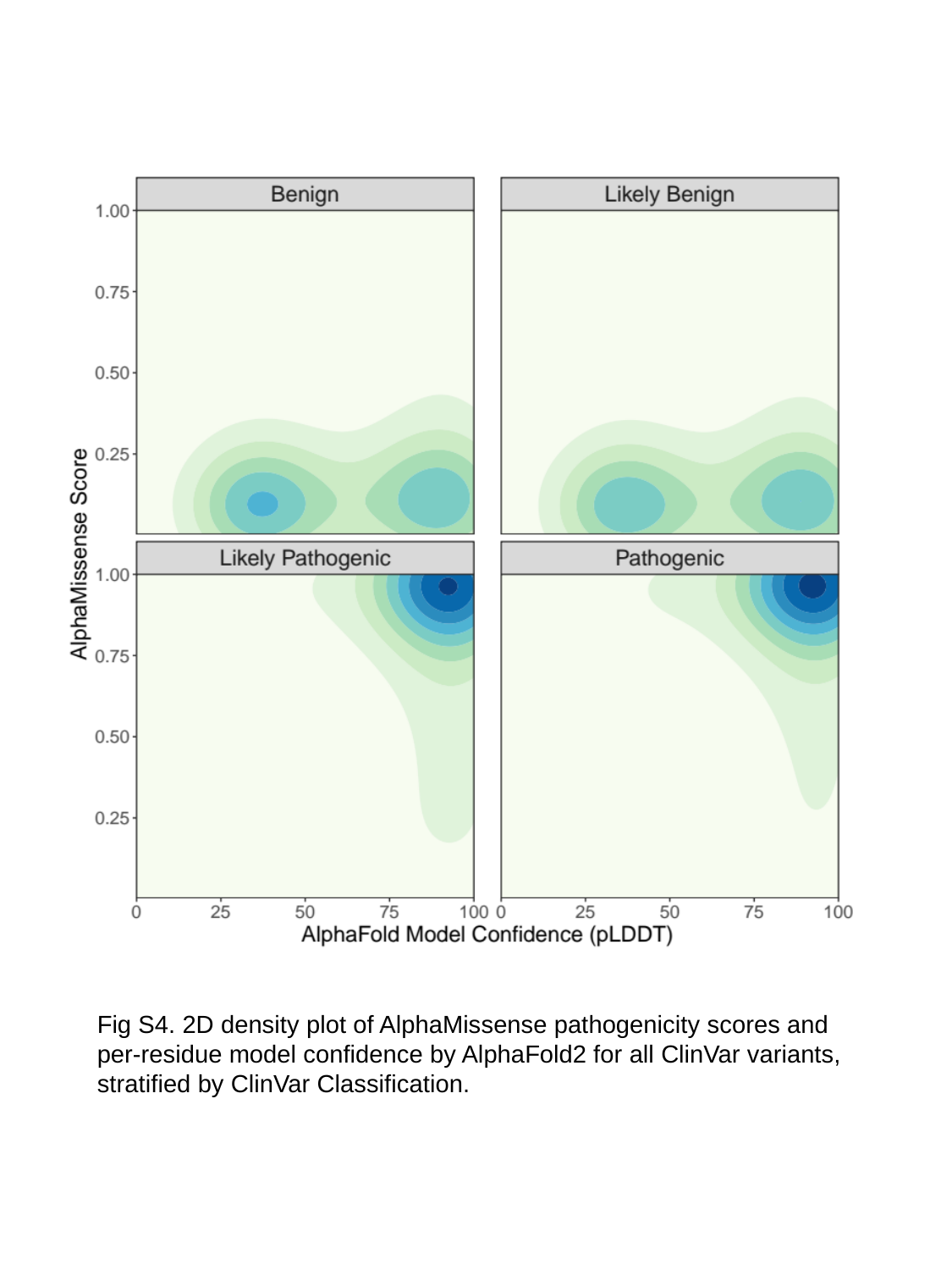

Fig S4. 2D density plot of AlphaMissense pathogenicity scores and per-residue model confidence by AlphaFold2 for all ClinVar variants, stratified by ClinVar Classification.
